# The COVID-19 pandemic sparked off a large-scale outbreak of carbapenem-resistant *Acinetobacter baumannii* from the endemic strains of an Italian hospital

**DOI:** 10.1101/2022.06.28.22276851

**Authors:** Greta Petazzoni, Greta Bellinzona, Cristina Merla, Marta Corbella, Ørjan Samuelsen, Jukka Corander, Davide Sassera, Stefano Gaiarsa, Patrizia Cambieri

**Affiliations:** Microbiology and Virology Unit. Fondazione IRCCS Policlinico San Matteo. Pavia, Italy; Department of Biology and Biotechnology. University of Pavia. Pavia, Italy; Norwegian National Advisory Unit on Detection of Antimicrobial Resistance, Department of Microbiology and Infection Control. University Hospital of North Norway. Tromsø, Norway; Institute of Pharmacy, Faculty of Health Sciences. UiT The Arctic University of Norway. Tromsø, Norway; Department of Biostatistics. University of Oslo. Oslo, Norway; Department of Mathematics and Statistics. University of Helsinki. Helsinki, Finland

## Abstract

*Acinetobacter baumannii* is a nosocomial pathogen that poses a serious threat due to the rise of incidence of multidrug resistant (MDR) strains. During the COVID-19 pandemic, MDR *A.baumannii* clones have caused several outbreaks worldwide. Here we describe a detailed investigation of an MDR *A. baumannii* outbreak that occurred at Fondazione IRCCS Policlinico San Matteo (Pavia, Italy). A total of 96 *A. baumannii* strains, isolated between January and July 2020 from 41 inpatients (both SARS-CoV-2 positive and negative) in different wards, were characterized by phenotypic and genomic analyses combining Illumina and Nanopore sequencing. Antibiotic susceptibility testing revealed that all isolates were resistant to carbapenems and the sequence analysis attributed this to the carbapenemase gene *bla*_OXA-23_. Screening of virulence factors unveiled that all strains carried determinants for biofilm formation, while plasmid analysis revealed the presence of two plasmids, one of which was a ⍰100kbp long and encoded a phage sequence.

A core genome-based phylogeny was inferred to integrate outbreak strain genomes with background genomes from public databases and from the local surveillance program. All strains belonged to the globally disseminated ST2 clone and were mainly divided into two clades. Isolates from the outbreak clustered with surveillance isolates from 2019, suggesting that the outbreak was caused by two strains that were already circulating in the hospital before the start of the pandemic. The intensive spread of *A. baumannii* in the hospital was enhanced by the extreme emergency situation of the first COVID-19 pandemic wave that resulted in minor attention to infection prevention and control practices.

**Importance:** The COVID-19 pandemic, especially during the first wave, posed a great challenge to the hospital management and generally promoted nosocomial pathogen dissemination. Multidrug resistant (MDR) *A. baumannii* can easily spread and persist for a long time on surfaces, causing outbreaks in healthcare settings. Infection prevention and control practices, epidemiological surveillance and microbiological screening are fundamental in order to control such outbreaks.

Here, we sequenced the genomes of 96 isolates from an outbreak of MDR *A. baumannii* strains using both short- and long-read technology in order to reconstruct the outbreak events in fine detail. The sequence data demonstrated that two endemic clones of MDR *A. baumannii* were the source of this large hospital outbreak during the first COVID-19 pandemic wave, confirming the effect of COVID-19 emergency disrupting the protection provided by the use of the standard prevention procedures.

## INTRODUCTION

*Acinetobacter baumannii* is an opportunistic gram-negative pathogen frequently associated with community-acquired and healthcare-associated infections (HAIs). This pathogen is a member of the ESKAPE group, despite causing only 2% of HAIs (1), due to its virulence and the substantial prevalence of antibiotic resistance.

Currently, the greatest cause of concern are strains that have shown to be rapidly capable of becoming resistant to a wide range of antibiotics (including front-line agents like carbapenems), thus becoming multidrug resistant (MDR). This is a critical issue in Southern Europe, especially in Italy, where the percentage of carbapenem-resistant *A. baumannii* (CRAB) strains during 2020 was 80.8% (WHO Regional Office for Europe/European Centre for Disease Prevention and Control. Antimicrobial resistance surveillance in Europe 2022 – 2020 data. Copenhagen: WHO Regional Office for Europe; 2022). In conjunction with antibiotic resistance, *A. baumannii* is concerning because of its exceptional capability to persist for long periods in the environment, even in the hospital setting, aided by biofilm production. This persistence on abiotic and biotic surfaces leads to chronic infections and facilitates its spread (2), especially in Intensive Care Units (ICUs) (3). Indeed, *A. baumannii* infections are typically related to medical equipment such as catheters and ventilators (i.e. pneumonia and bloodstream infections), while urinary infections and those affecting skin, soft tissues and surgical sites are less common (4). In light of this, the World Health Organization included MDR *A. baumannii* in the critical group, a list of bacteria that pose the greatest threat to human health and for which the development of novel antibiotics is urgently needed (5).

The COVID-19 pandemic has been posing an arduous challenge for hospitals worldwide, especially in the early period. The increasing COVID-19-related hospitalizations led to shortages in personnel, personal protective equipment and medical equipment. These, in turn, often resulted in the impossibility to maintain strict infection prevention and control (IPC) practices and consequently in an increase of bacterial and fungal infections. In fact, several *A. baumannii* outbreaks have been described during the first pandemic wave worldwide; e.g. (6,7). Fondazione IRCCS Policlinico San Matteo (Pavia, Italy) played a key role in the management of COVID-19 pandemic in North Italy, especially during the first wave, being one of the largest hospitals (⍰900 beds) in the Lombardy region. An active genomic surveillance programme and IPC containment strategies for ESKAPE pathogens were already in place in the hospital before COVID-19. Thanks to surveillance measures, the prevalence of *A. baumannii* in the 2018-2019 biennium was less than one isolate per 1000 days of hospitalizations. However, during the first wave of COVID-19 a large outbreak of MDR *A. baumannii* emerged. Here we report a detailed genomic characterization of this outbreak.

## MATERIALS AND METHODS

### Data collection

This retrospective study was conducted at IRCCS Fondazione Policlinico San Matteo in Pavia (Italy), where 151 isolates of *A. baumannii* were collected from 06 January, 2020 to 30 July, 2020. 96 isolates were chosen to conduct this study; the choice was made favoring the inclusion of both samples from colonization and infection materials for each patient (where available) and of multiple samples in case of long hospitalizations (1 sample/7 days/material). The isolates selected were identified with MALDI–TOF mass spectrometry (Bruker Daltonik, Bremen, Germany) equipped with Bruker biotyper 3.1 software. Antibiotic susceptibility was tested with both Sensititre™ Gram Negative EUMDROXF Plate (ThermoFisher Scientific, Rodano, Italy) and Phoenix™ NMIC/402 panel loaded on BD Phoenix™ M50 instrument. Values were interpreted according to the European Committee on Antimicrobial Susceptibility Testing (EUCAST) clinical breakpoints (vr. 10). Tigecycline was interpreted following Clinical Laboratory Standard Institute (CLSI) breakpoints (30^th^ ed.).

Patient information was retrieved from the hospital database. COVID-19 positivity was defined at the time of admission, consulting the results of real-time reverse transcriptase PCR tests for the presence of SARS-CoV-2.

### DNA extraction and genome characterization

The genomic DNA of the 96 strains was extracted with DNeasy Blood & Tissue Kit (QIAGEN) for short-read sequencing using an Illumina platform and with MagAttract HMW DNA Kit (QIAGEN) for long-read sequencing using Nanopore technology. Short and long reads were combined with the Unicycler software (8) to perform hybrid assemblies. The genomes obtained were then *in silico* sequence typed (ST) using the Pasteur MLST scheme (9) with an in-house script. Presence of virulence and resistance genes was assessed using ABRicate [https://github.com/tseemann/abricate] with the NCBI AMRFinderPlus (10), CARD (11), Resfinder (12), and VirulenceFinder (13) databases. The presence of insertion sequences was determined with MobileElementFinder (14).

The presence of plasmids was evaluated with PlasmidFinder (15) and a more in-depth plasmid search was performed by visualizing the assembly graph in Bandage (16) for the least fragmented isolates, checking for circularity and higher depth coverage. The presence of the plasmids identified was assessed in all the remaining assemblies using blastn. Plasmids were annotated combining Prokka (17), NCBI Conserved Domain Database (18) and possibly blastp on the nr database. PHASTER (19) was then used to identify and annotate prophage sequences. In order to understand the prevalence of the plasmids found with our approach in the *A. baumannii* overall population, we created a collection of *A. baumannii* genomes joining those from this study and all high-quality genomes from the PATRIC DB (sequences were retrieved in July 2022, using the *makepdordb*.*py* script of the P-DOR pipeline [https://github.com/SteMIDIfactory/P-DOR]). All assemblies were then scanned for the presence of all plasmid genes; genomes were considered to carry a plasmid when they encoded the vast majority of its genes (≥11 genes for the ⍰8 kbp; ≥120 genes for the ⍰100 kbp). Finally, FastTree (20) was used to infer the global phylogeny of the species, thus allowing to study the distribution of the plasmids among the population.

### Surveillance program genomes

Strains selected by the genomic surveillance program of the hospital are routinely processed for short-read sequencing as follows: DNA is extracted using DNeasy Blood & Tissue Kit (QIAGEN), sequenced with an Illumina platform and assembled using Unicycler (8) with default parameters. We retrieved all the ST2 genomes obtained by the surveillance program (N = 23) in the months before and after the outbreak period (12 from 2019, five from 2020 and six from 2021) and we added them to our dataset.

### Phylogenetic analyses

In order to characterize the outbreak of *A. baumannii* a genomic background was constructed. In detail, the twenty high-quality genomes most similar to those in our set according to k-mer content similarity (MASH (21)) were retrieved from the PATRIC DB [https://www.patricbrc.org]. Then, the P-DOR pipeline [https://github.com/SteMIDIfactory/P-DOR; version beta1.5] was used to align all genomes in our dataset to an internal reference (the complete genome of strain 4946) and to extract coreSNPs. Maximum Likelihood phylogeny was inferred with RAxML (22) (using 100 bootstraps resamples) on the resulting coreSNP alignment using the general time reversible model, as suggested by ModelTest-NG (23), with ascertainment bias correction (24).

## RESULTS

### Dataset characterization

During the first COVID-19 wave an increase of incidence of *A. baumannii* strains was registered in the study hospital; the cases raised from a baseline of less than one isolate per 1000 days of hospitalization in the 2018-2019 biennium to 2.8 isolates. We characterized 96 strains collected in this period (from January to July 2020) from 41 individuals hospitalized in the ICUs (N = 37), Pneumology (N = 2) and Otorhinolaryngology (N=1) wards. Three samples were collected from one patient, who was hospitalized in another care institute in Pavia (2 samples) and later admitted to the Emergency department of the study hospital (1 sample). Patients had a mean age of 61 (SD: +/- 10 years), ranging from 30 to 83 years old and were predominantly male (87.8%; N = 36). The median length of stay was 47 days (interquartile range [IQR] = 25-74 days) and 43.9% (N = 18) of patients died (median length of stay = 52 days; IQR = 37-85 days). Thirty patients tested positive and seven negative to SARS-CoV-2; the remaining four were hospitalized before the pandemic period. 33.3% (N = 32) of the *A. baumannii* isolates were related to lower respiratory tract infections and 22.9% (N = 22) to bloodstream infections; 3.1% of strains were isolated from the urinary tract (N=3) and the other 40.6% from rectal swabs (N = 38) and nasal swab (N = 1). Supplementary table 1 summarizes isolate and patient information.

All 96 isolates were sequenced using both long- and short-read technology. The hybrid assemblies enabled us to obtain eight complete and 87 high-quality genomes (mean length of the genomes = 4 Mbp; mean number of contigs = 11.3, Supplementary table 2). The remaining one was discarded (sample 4964) and excluded from all downstream analyses. Genome analysis revealed that all 95 isolates belonged to ST2, which is part of the globally disseminated International Clone II (IC2) (25).

### Phylogenetic analysis

In order to further characterize the outbreak, we performed a phylogenetic analysis using the 95 *A. baumannii* genomes from the outbreak, 23 from the hospital surveillance program, and 367 from the PATRIC database as background. The inferred phylogeny (Figure 1) depicts a situation in which the *A. baumannii* outbreak in the hospital was caused by multiple strains. The majority of the outbreak isolates (N = 93) are divided into two monophyletic clades: one larger (cluster 1, 71 outbreak strains in a clade of 80 total genomes) and one smaller (cluster 2, 23 outbreak strains). Cluster 1 also includes nine isolates from the hospital surveillance program (one from 2019, four from 2020 and four from 2021) and one from the PATRIC DB. Cluster 2 is exclusively composed of 2020 isolates; nonetheless, isolate 4200_2019 from 2019 is basal to this cluster. A background isolate from Belgium (470.13595 of PATRIC DB) interposes between the 2019 surveillance isolate and cluster 2; however the common branch to these genomes has very limited support (29/100 bootstrap). Interestingly, the genome of one of the two remaining isolates from January 2020 (4636_2020) clustered with four 2019 surveillance genomes, while the second one (4614_2020) clustered with one surveillance genome from late 2020.

**Figure 1.**
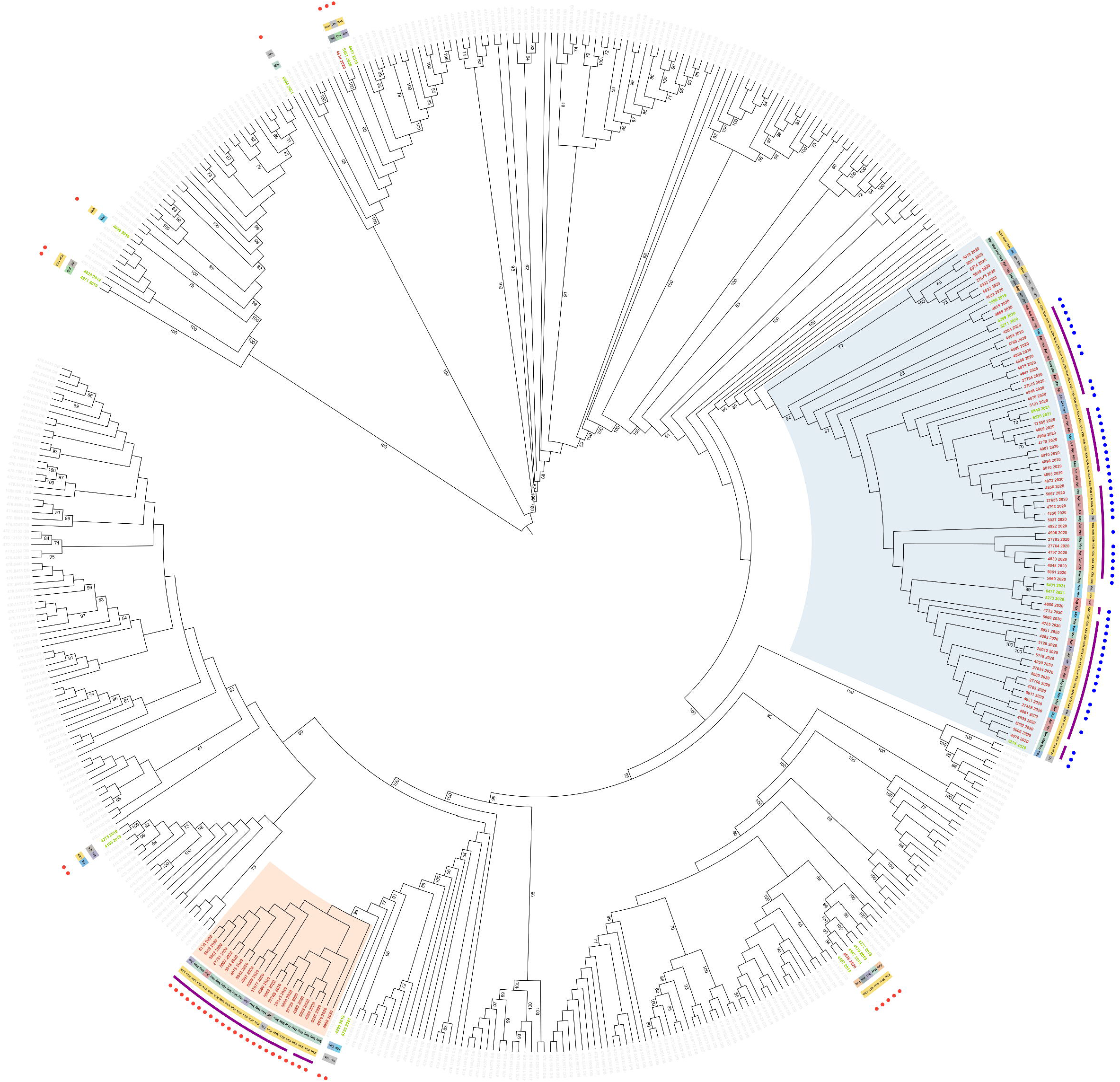
Maximum likelihood phylogeny of 485 *A. baumannii* strains: 95 outbreak genomes (red labels), 23 surveillance genomes (green labels) and 367 PATRIC genomes (gray labels) inferred on coreSNPs with RAxML. Date of isolation (innermost crown), ward (second crown), and SARS-CoV-2 positivity (third crown) were retrieved from the hospital and microbiological database. The presence of 100 kb plasmid (blue dots) and *pgaC* gene (red dots) were determined by genomic analyses. The two main outbreak clusters are highlighted in blue (Cluster 1) and red (Cluster 2). Bootstrap values above 50/100 are indicated on tree branches. This figure was obtained using iTol [https://itol.embl.de/].

### Antibiotic resistance and resistance/virulence genes

All 95 outbreak isolates were resistant to carbapenems (100% to imipenem and meropenem). Almost all isolates presented resistance to aminoglycosides: 97.9% (N = 93) were found to be resistant to gentamicin and 98.9% (N = 94) to amikacin. Moreover, only 8.9% (N = 7) of isolates tested were resistant to tigecycline and eight isolates were susceptible to increased exposure to this antibiotic. Additionally, the totality of isolates tested for the ciprofloxacin (N = 90) were resistant. None of the isolates was found resistant to colistin. The antibiotic resistance factors predicted in the genomes were concordant with the phenotypes. Resistance to carbapenems was confirmed by the presence of *bla*_OXA-23_. The intrinsic *bla*_OXA-51_-variant, *bla*_OXA-66_ was also detected but no ISAba1 sequence insertion was found upstream. All strains carried *bla*_ADC-25_, the majority (N = 89) also presented *bla*_ADC-73_, other five *bla*_ADC-56_ and the remaining one had none of these two (Supplementary table 3).

Virulence determinants were also found in outbreak isolates, particularly those involved in biofilm formation. In detail, all isolates presented the *ompA* gene which is important for the development of robust biofilms on abiotic surfaces (26) and the *bap* gene which ensures a strong biofilm formation (27). 94 out of 95 isolates had *csuAB, csuA, csuB, csuC, csuD* and *csuE* that encode for the Chaperone–Usher secretion (CUS) system, fundamental for the first bacterial attachment on abiotic surfaces (28). The sole strain not carrying the CUS system was isolated in January 2020 (4614_2020). Strains also had genes for the BfmS/BfmR, a two-component system that plays a key role in CUS systems regulation and *ompA (29)*. Finally, 23.3% (N = 25) isolates presented a cluster of four genes (*pgaA, pgaB, pgaC*, and *pgaD*) encoding for the Poly-β-(1, 6)-N-Acetlyglucosamine (PNAG), a component involved in biofilm development (30). Interestingly, these isolates included two from January 2020 (4636_2020, 4614_2020) and those in cluster 1. In the remaining outbreak isolates (N = 70) the *pcaC* gene was interrupted by the ISAba1 insertion sequence (Supplementary table 4).

### Plasmids

We investigated the presence of plasmids in the outbreak genomes and found that all the samples shared a plasmid of 8731 bp which perfectly matches with NZ_CP008708.1, containing twelve coding sequences, ten being hypothetical proteins. Most of the genomes of cluster 1 (51/71) presented also a second 106175 bp long plasmid (highly similar to CP081146.1). Neither of the two plasmids encoded for virulence or resistance genes. We analyzed the ⍰100 kbp plasmid in-depth; 83 out of its 131 coding sequences were annotated as hypothetical, 20 of which reported to be phage-associated. Indeed, the majority of the plasmid sequence consisted in a profagic region of ⍰85 kbp (9261-95595). The phylogeny in Figure 1 indicates that this plasmid is present only in cluster 1. Genomes in the basal position of the cluster do not carry the plasmid, suggesting that it was acquired with a single transfer event, at the beginning of the outbreak. Also, the pattern of plasmid presence within the cluster indicates multiple losses. Lastly, we investigated the presence of the ⍰8 kbp and the ⍰100 kbp plasmids among the overall *A. baumannii* population (N = 8969 genomes) and found them to be differentially distributed (Supplementary figure 1). The first one was present in the 32.1% (N = 2882), instead the second one in the 7% (N = 636) of the population.

## DISCUSSION

In this work we present the data from a hospital-wide outbreak of MDR *A. baumannii* that occurred during the first wave of the COVID-19 pandemic. A rise in the incidence of HAIs during COVID-19 pandemic waves was observed in the study hospital and also in other hospitals and countries worldwide (31, 32 33). This is particularly true for *A. baumannii*, which was the most frequently reported cause of hospital outbreaks, together with methicillin resistant *Staphylococcus aureus* and fungal infections (34), (35), (36). For example, during the first wave, Perez et al. reported a hospital-acquired carbapenem-resistant *A. baumannii* (CRAB) outbreak of 34 isolates harboring *bla*_OXA-23_ which involved 34 patients admitted to ICUs for COVID-19 patients in New Jersey hospital (6). Also Gottesman and colleagues (7) reported an outbreak in Israel of five CRAB carrying *bla*_OXA-24_ and belonging to IC2 in ICUs patients.

In our study, we were able to identify two large phylogenetic clusters (Figure 1), which included most of the outbreak strains. The inclusion of genomes sequenced before and after the outbreak by the hospital surveillance program revealed that multiple strains were circulating in the hospital before the start of the pandemic, including the two (3996_2019 and 4200_2019) that caused the outbreak.

We detected two plasmids within our genomes: one of ⍰8 kbp and another of ⍰100 kbp. The second one is specific to a subclade of cluster 1 and contains a large prophage sequence carrying mostly uncharacterized genes. Four strains (4682_2020, 3996_2019,4615_2020,4669_2020), which are basal to the subclade, were isolated in 2019 and early 2020 and their genomes did not contain the ⍰100 kbp plasmid. This suggests that the plasmid was acquired later during the outbreak, possibly within the hospital. The hypothesis is reinforced by the presence of another subclade of this cluster (N genomes = 8) in which the plasmid is absent. Moreover, this phage-encoding plasmid is supposedly specific to *A. baumannii* as it is not present in other species (Blastn on the nt database performed on 4 October 2022). This observation suggests that the source of acquisition could have been another strain of *A. baumannii*, not detected by our surveillance program. Despite its fitness advantage not being clear, we can observe a tendency to preserve the plasmid in the genomes analyzed. This trend can be observed both in the local phylogeny (Figure 1) and in the global one (Supplementary figure 1).

Mapping patient information on the tree revealed that SARS-CoV-2 coinfection was approximately equally distributed between the two phylogenetic clusters. Thus, no association between *A. baumannii* clusters and viral infection can be hypothesized. The possibility of transmission of *A. baumannii* between COVID-19 positive and negative patients, in absence of viral co-transmission is interesting, and can be explained by the high capability of *A. baumannii* to survive on abiotic surfaces due to biofilm formation (37). In turn, this could also explain why this bacterial species thrived during the emergency period, both in the study hospital and in other institutions worldwide. Moreover no association was found between clusters and wards, outcome, or age of patients.

The phylogeny we inferred allowed us to identify eight separate clusters of study genomes (including the two large ones), four of which contained strains isolated months before and after the epidemic event. This observation unveils the presence of at least four endemic clones in the study hospital, two of which gave origin to the large outbreak described in this work. Interestingly, no genomic factor that is known to enhance bacterial spread was found in the study genomes. This result indicates that the spread of the bacterial pathogen was enhanced solely by the extreme emergency situation caused by the first wave of the COVID-19 pandemic. Personnel and bed shortage, and the prioritization of self-preservation from the viral infection of healthcare workers, led to unavoidable diminished attention towards the standard patient handling measures (e.g. changing gloves between patients). In accordance with this hypothesis, the incidence of *A. baumannii* decreased to lower than one isolate per 1000 days of hospitalizations after the first pandemic wave and remained under control during the following waves, thanks to a better management of COVID-19 patients and to a better knowledge of the viral disease.

## Ethical statement

The study was designed and conducted in accordance with the Helsinki declaration and approved by the Ethics Committee of Fondazione IRCCS Policlinico San Matteo in Pavia, Italy (internal project code: 08022298/13, Project number: 733-rcr2013-34). The work described herein is a retrospective study performed on bacterial isolates from human samples that were obtained as part of routine hospital care.

## Data Availability

All data produced in the present study are available upon reasonable request to the authors

## Funding section

J.C. was supported by the ERC grant 742158. S.G. was supported by Ricerca Corrente grant 924-rcr2018i-34 (Italian Ministry of Health)

## Acknowledgement section

We acknowledge Anna K. Pöntinen and Francois Cleon for experimental assistance and the Genomics Support Centre Tromsø, UiT The Arctic University of Norway for the short- and long-read sequencing.

## Figure Legends

Supplementary figure 1- Global phylogeny of 8969 strains of *A. baumannii*. The presence of 8 kb plasmid (green labels) and 100 kb plasmid (red labels) were determined by genomic analyses. This figure was obtained using iTol [https://itol.embl.de/].

## Table Titles

Supplementary table 1 - Isolates of *A. baumannii* information and patient metadata.

Supplementary table 2 - Characteristics of the 96 *A. baumannii* genomes.

Supplementary table 3 - Phenotypic resistance and presence of antibiotic resistance genes in the 95 *A. baumannii* isolates.

Supplementary table 4 - Presence of virulence genes in the 95 *A. baumannii* isolates.

