## Supplementary figures and images for "The COVID-19 pandemic sparked off a large-scale outbreak of carbapenem-resistant *Acinetobacter baumannii* from the endemic strains of an Italian hospital"

### Supplementary Figure 1

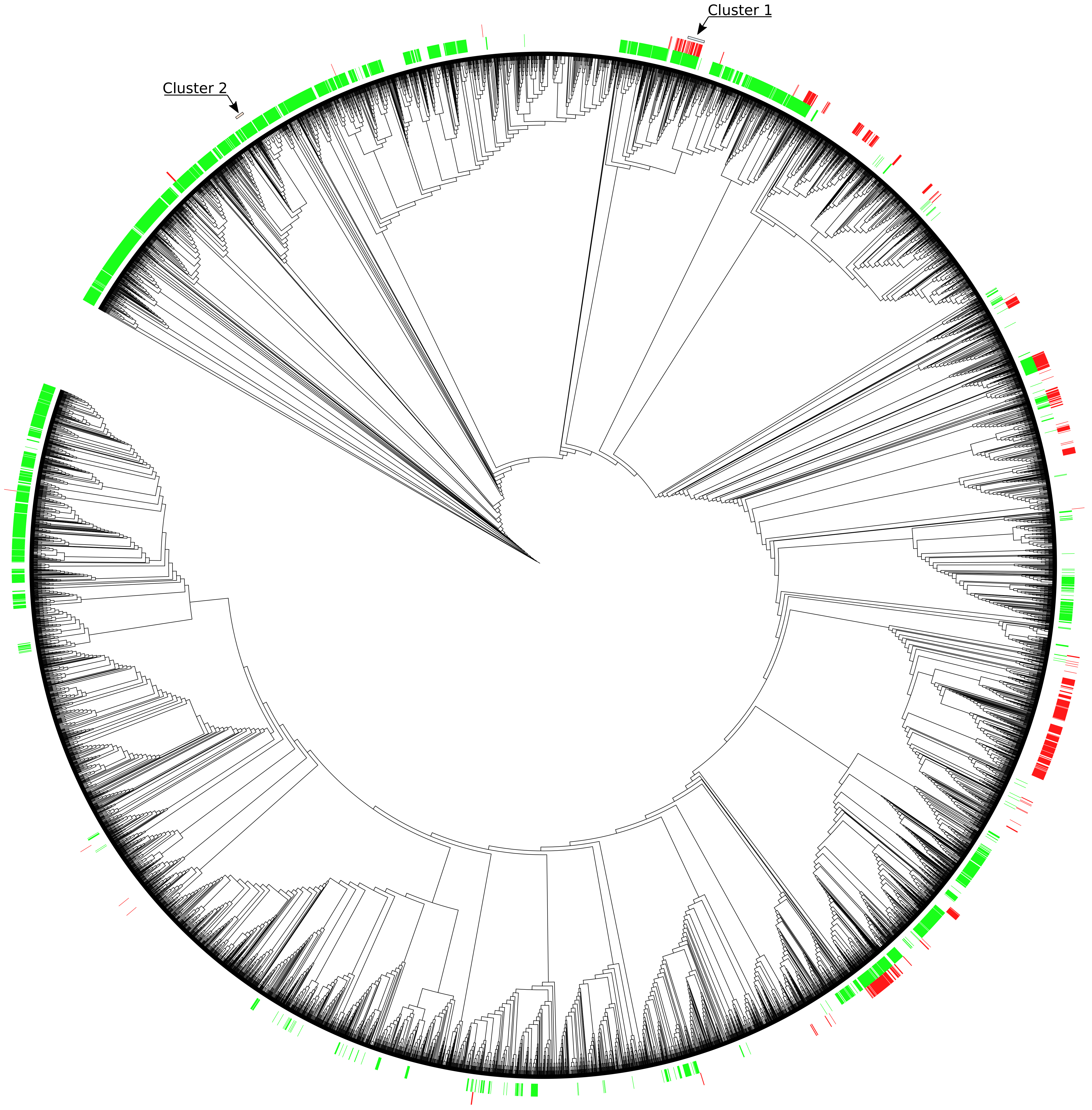
